# PSMA+ Extracellular Vesicles are a Biomarker for SABR in Oligorecurrent Prostate Cancer Analysis from the STOMP-like and ORIOLE trial cohorts

**DOI:** 10.1101/2024.09.14.24313680

**Authors:** Jack Andrews, Yohan Kim, Edlira Horjeti, Ali Arafa, Heather Gunn, Aurélie De Bruycker, Ryan Phillips, Daniel Song, Daniel S. Childs, Oliver A. Sartor, Jacob J. Orme, Aadel A. Chaudhuri, Phuoc Tran, Ana Kiess, Philip Sutera, Carole Mercier, Piet Ost, Sean S. Park, Fabrice Lucien

**Author notes:** Equal contribution (Phuoc Tran, Ana Kiess, Philip Sutera, Carole Mercier, Piet Ost, Sean S. Park, Fabrice Lucien). Correspondence **Fabrice Lucien, PhD** Kellen Building 3-121 Department of Urology, Mayo Clinic, Rochester MN, USA Department of Immunology, Mayo Clinic, Rochester MN, USA **Sean S. Park, MD PhD** Charlton Building SL-121 Department of Radiation Oncology, Mayo Clinic, Rochester MN, USA.

## Abstract

**Purpose:** Two randomized clinical trials (STOMP and ORIOLE) demonstrated that stereotactic ablative radiotherapy (SABR) can prolong ADT-free survival or progression-free survival (PFS) in patients with metachronous oligometastatic prostate cancer (omCSPC) patients. While most omCSPC patients have a more modest delay in progression, a small subset achieves a durable response following SABR. We investigated the prognostic and predictive value of circulating PSMA-positive extracellular vesicles (PSMA^+^EV) and prostate specific antigen (PSA) in a biomarker correlative study using blood samples from three independent patient cohorts.

**Methods:** Plasma samples from 46 patients on the ORIOLE trial and 127 patietns on the STOMP trial protocol with omCSPC patients treated with SABR. Pre-SABR PSMA^+^EV levels (EVs/ml) were measured by nanoscale flow cytometry. Kaplan-Meier curves and logistic regression models were used to determine the association of PSMA^+^EV and PSA levels with clinical outcomes.

**Results:** In the pooled cohorts, median bPFS were 26.1 and 15.0 months (p=0.005) and median rPFS were 36.0 and 25.0 months (p=0.003) for PSMA^+^EV low and high groups, respectively. The combination of pre-SABR low levels of both PSMA^+^EV and PSA was associated with lower risk of radiographic progression (HR=0.34, 95% CI: 0.18-0.58, p=0.0002). In the ORIOLE cohort, which included both a SABR arm and an observation arm, low PSMA^+^EV was predictive of benefit from SABR (p=0.012).

**Conclusions:** PSMA^+^EV is a novel prognostic and predictive biomarker of radiographically occult tumor burden in omCSPC. PSMA+EV may inform clinical decisions regarding which patients achieve a durable benefit from consolidative SABR alone.

## Introduction

The concept of oligometastatic disease was first suggested in 1995 by Hellman and Weichselbaum as an unique biologic state between localized and widespread metastatic disease.^1^ This concept has changed the dogma, now proposing that patients with early metastatic disease may still be cured with a combination of local and metastasis-directed therapies (MDT). SABR-COMET was the first trial to evaluate stereotactic body radiotherapy (SABR) in various oligometastatic cancers.^2^ This trial was also the first to demonstrate a survival benefit with SABR, however prostate cancer only represented 16% of the treatment group. In prostate cancer, two randomized clinical trials have since demonstrated an oncologic benefit of SABR in the oligometastatic setting (STOMP and ORIOLE).^3,4^

SABR is rapidly emerging as a safe and potentially curative treatment option in omCSPC with the potential to delay or avoid the use of non-curative systemic therapy, delaying the negative quality of life impact of androgen deprivation and the development of castration refractory disease. Both the STOMP and ORIOLE trials reported long-term disease free survival in ∼20% of patients, however most patients progressed and ultimately received systemic therapy.^3,4^ This divergence in outcomes suggests that selected omCSPC patients can benefit from SABR alone, while may need earlier systemic therapy. This highlights a current unmet need to develop predictive biomarkers that can help select patients who may attain a durable response from SABR alone without systemic therapy. Unfortunately, to date no such effective liquid biomarkers exist.

Tumor-derived extracellular vesicles (tdEVs) are microscopic particles released by tumors into the bloodstream and are emerging as non-invasive biomarkers for cancer detection and prognosis.^5^ An important benefit of circulating tdEVs is that, unlike circulating tumor cells and circulating tumor DNA, they are detectable across the entire spectrum of the disease from localized prostate cancer to widely spread metastatic castration resistant prostate cancer (CRPC).^6,7^ We recently demonstrated the positive association of circulating levels of PSMA^+^EV with radiographic tumor burden in prostate cancer.^6^ We also reported that pre-SABR PSMA^+^EV can predict risk of disease recurrence in omCRPC patients treated with SABR. Herein, we conducted a biomarker correlative study evaluating the prognostic and predictive value of PSMA^+^EV in a multi-institutional cohort of omCSPC patients.

## Materials and Methods

### Patient Cohorts

Three cohorts of patients with stored plasma available were obtained from Ghent University (PI: Piet Ost, Belgium) (N=80), the Iridium Network (PI: Carole Mercier, Belgium) (N=47) and the ORIOLE trial (PI: Phuoc Tran) (N=46)^3^. Study approval was granted by the Mayo Clinic Institutional Review Board (IRB #21-004451). The Ghent University cohort enrolled patients between 2015 and 2020 under the existing STOMP trial protocol (PI: Piet Ost).^4^ In the Iridium Network cohort, patients were treated for omCSPC with SABR between 2018 and 2020. Detailed characteristics of the ORIOLE trial has been previously reported.^3^

### Enumeration of PSMA-Positive Extracellular Vesicles (PSMA^+^EV)

Platelet-poor plasma (PPP) samples were incubated with fluorescent PSMA antibodies (J591 clone) and concentrations of PSMA^+^EV were measured by nanoscale flow cytometry as previously described.^6,8^ All measurements were performed blinded from clinical data. Each sample was run in three technical replicates and average was used for data analysis. Following standardized reporting of EV flow cytometry experiments, a detailed description of the methodology can be found in the MIFlowCyt-EV report (**Supplementary Materials**).

### Statistical Analysis

Biochemical progression-free survival (bPFS) and radiographic progression-free survival (rPFS) were used as clinical endpoints to determine the association of PSMA^+^EV levels, serum PSA and PSA doubling time (PSA DT) at 3 months with oncological outcomes. Biochemical progression was defined as any of the following: i) for patients who have undergone a radical prostatectomy, PSA rise to ≥ 0.2 ng/mL from nadir after MDT; or ii) for patients who did not experience a PSA nadir below 0.2 ng/mL after MDT: then first rise in PSA after reaching nadir or iii) for patients treated with primary radiation to the prostate, PSA nadir +2 ng/mL after MDT or iv) initiation of systemic therapy; local recurrence, or distant recurrence prior to reaching numerical definition of PSA as above. Radiographic progression was defined as new nodal lesions, intrapelvic or distant, bone, or visceral lesions on conventional (bone scintigraphy) or molecular (C-11 Choline PET/CT or PSMA-PET CT) imaging with application of the RECIST version 1.1 criteria. Kaplan-Meier (KM) estimates were used to estimate survival curves. For each KM plot, p values were derived from the log-rank test for difference between groups. For association of PSMA^+^EV and PSA with oncological outcomes, concentrations were converted to categorical variables and patients were classified as high and low. The optimal cutoff values of PSMA^+^EVs and PSA were defined as the value with the most significant log-rank test split in univariate Cox proportional hazard model.^9^ The hazard ratio (HR) of each biomarker was calculated with each clinical outcome (bPFS and rPFS). We tested the effect of PSMA^+^EVs on bPFS and rPFS controlling for the effect of PSA levels, number of lesions, and lesion location in mutivariate Cox proportional hazard models. PSA DT was also treated as categorical variable (≤ or > 3 months). For predictive biomarker assessment, we tested if PSMA+EV was predictive by including PSMA+EV, treatment group, and treatment-by-biomarker interaction term in a Cox proportional hazard model. Hazard ratios (HRs) with 95% confidence intervals were calculated. Statistical software (SAS, version 9.4, SAS Institute Inc, Cary, NC) was used for the multivariate Cox models. Prism v9.0.1 (GraphPad Software) was used for all other statistical analyses.

## Results

### Baseline Characteristics, PSMA^+^EV levels and Survival Outcomes

One hundred and seventy-three patients with plasma samples available were included (Ghent University n=80, Iridium Network n=47, ORIOLE Trial n=46). The diagram for the study cohorts is included in **Supplementary Figure S1.** Sixteen patients with CRPC disease or local recurrence/non-metastatic disease were excluded from the study. For the ORIOLE cohort, 30 patients were treated with SABR and 16 patients were part of the observation arm. Patient characteristics for the three cohorts are presented in **Table 1**. For all patients treated with SABR (n=157), no active systemic therapies were received concomitantly with SABR. Per ESTRO-EORTC classification^10^, 92% (145/157) of patients were diagnosed with metachronous oligorecurrent prostate cancer. Oligometastatic disease was diagnosed with advanced PET imaging (80%) and conventional imaging (20%). The median follow up was 45.7 months (range 41.2-51.7). Median bPFS was 21.5 months in the Ghent University cohort, 17.5 months in the Iridium cohort and 11.1 months in ORIOLE SABR arm (**Supplementary Figure S2A)**. Median rPFS was 29.0 months in the Ghent University cohort, 32.1 months in the Iridium cohort and 25.3 months in ORIOLE SABR arm (**Supplementary Figure S2B)**.

**Table 1.** Patient Characteristics.

|  | **Ghent University** | **Iridium Network** | **ORIOLE** |
| --- | --- | --- | --- |
| **Number of patients** | 80 | 47 | 46 |
| **ESRO-EORTC classification** |  |  |  |
| Metachronous oligorecurrence | 80 (100%) | 35 (74%) | 46 (100%) |
| Repeat oligorecurrence | 0 (0%) | 7 (15%) | 0 (0%) |
| Synchronous | 0 (0%) | 5 (11%) | 0 (0%) |
| **Treated with SABR** |  |  |  |
| No | 0 (0%) | 0 (0%) | 16 (35%) |
| Yes | 80 (100%) | 47 (100%) | 30 (65%) |
| **Prior ADT** |  |  |  |
| No | 80 (100%) | 18 (38%) | 25 (54%) |
| Yes | 0 (0%) | 29 (62%) | 21 (46%) |
| **PSA at MDT** |  |  |  |
| Median | 1.9 | 2 | 6.8 |
| Min | 0.21 | 0.02 | 0.4 |
| Max | 72.1 | 130.4 | 33 |
| **PSA Doubling Time** |  |  |  |
| < 3 months | 18 (23%) | 15 (32%) | 8 (17%) |
| ≥ 3 months | 57 (71%) | 25 (53%) | 38 (83%) |
| Unknown | 5 (6%) | 7 (15%) | 0 (0%) |
| **ADT at MDT** |  |  |  |
| No | 76 (95%) | 47 (100%) | 46 (100%) |
| Yes | 4 (5%) | 0 (0%) | 0 (0%) |
| **Type of Imaging** |  |  |  |
| Bone scan | 0 (0%) | 2 (4%) | 46 (100%) |
| Choline PET | 80 (100%) | 0 (0%) | 0 (0%) |
| PSMA PET | 0 (0%) | 44 (94%) | 0 (0%) |
| FDG PET | 0 (0%) | 1 (2%) | 0 (0%) |
| **Number of lesions treated** |  |  |  |
| 1 | 40 (50%) | 28 (60%) | 19 (41%) |
| 2 | 19 (24%) | 14 (30%) | 13 (28%) |
| 3 | 12 (15%) | 4 (8%) | 14 (31%) |
| >3 | 9 (11%) | 1 (2%) | 0 (0%) |
| **Sites of lesions** |  |  |  |
| Node only | 49 (61%) | 23 (49%) | 27 (59%) |
| Bone only | 25 (31%) | 18 (38%) | 10 (22%) |
| Multi-site | 6 (8%) | 6 (13%) | 9 (19%) |
| **Median Follow-up (months)** | 51.7 | 43.8 | 41.2 |
| **PSMA+EV (EVs per ml)** |  |  |  |
| Median | 5.91E+06 | 4.98E+06 | 2.12E+06 |
| Min | 1.42E+06 | 1.60E+06 | 3.33E+05 |
| Max | 4.44E+07 | 1.79E+07 | 5.79E+08 |

**Table 2.** Risk of disease progression for baseline PSA and PSMA+EV levels in the pooled STOMP-like and ORIOLE cohort.

|  |  |  | Number of patients | Hazard Ratio | 95% CI | P value |
| --- | --- | --- | --- | --- | --- | --- |
| Pooled cohort | bPFS | PSA low | 62 | 0.66 | 0.43-1.01 | 0.053 |
|  |  | PSMA-EV low | 68 | 0.6 | 0.40-0.92 | 0.018 |
|  |  | **PSA low +PSMA-EV low** | **29** | **0.36** | **0.19-0.67** | **0.001** |
|  | rPFS | PSA low | 62 | 0.54 | 0.31-0.82 | 0.009 |
|  |  | PSMA-EV low | 68 | 0.57 | 0.36-0.90 | 0.02 |
|  |  | **PSA low +PSMA-EV low** | **29** | **0.27** | **0.12-0.58** | **0.0009** |

Comparative analysis between baseline (pre-SABR) PSA levels demonstrates no significant difference between the Ghent University and Iridium Network cohorts (1.9 and 2.0 ng/ml, p>0.99) **(Supplementary Figure S3A)**. There was a significant difference in baseline PSA when comparing the Ghent University and Iridium Network cohorts (p<0.0001) with the ORIOLE cohort (6.8 ng/ml) which could be explained by the lower sensitivity of conventional imaging (ORIOLE) over PET imaging (both Belgium centers) for detecting radiographic recurrence. Comparative analysis between baseline PSMA^+^EVs demonstrate no significant difference between the Ghent University and Iridium Network cohorts (5.91x10^6^ and 4.98x10^6^ EVs/ml, p=0.74). Similar to baseline PSA, there was a significant difference in baseline levels of PSMA^+^EVs in the ORIOLE cohort (median: 2.12x10^6^ EVs/ml) when compared to the Ghent University (p<0.0001) and Iridium Network (p=0.0004) cohorts (**Supplementary Figure S3B).** No correlation was found between baseline levels of PSA and PSMA^+^EVs in the three cohorts (**Supplementary Figure S2C)**. Given the similarities in patient characteristics, treatment plan and oncological outcomes of the Ghent University and Iridium Network cohorts with the STOMP trial^4^, we combined both cohorts and referred to them as the “STOMP-like” cohort hereafter. Oncological outcomes of patients stratified by the number of metastatic lesions can be found in **Supplementary Figure S4.**

### Baseline PSMA^+^EV is a prognostic biomarker in oligorecurrent castration-sensitive prostate cancer

Following stratification of patients based on PSMA^+^EV levels, low and high baseline levels of PSMA^+^EVs in the pooled cohort of the ORIOLE trial and STOMP-like Cohorts, a median bPFS were 26.1 and 15.0 months (p=0.005) (**Figure 1A**). Median rPFS were 36.0 and 25.0 months for patients with low and high PSMA^+^EVs (p=0.003) (**Figure 1B**). Low baseline levels of PSMA^+^EVs was associated with lower risk of bPFS (HR=0.59, 95% CI: 0.40-0.85, p=0.005) and rPFS (HR=0.55, 95% CI: 0.34-0.81, p=0.003). Patients stratified based on PSA and PSA doubling time (PSA DT) demonstrated similar differences in PFS outcomes, but baseline PSA was a superior predictor of outcome compared to PSA DT (**Supplementary Figures S5 – S7)**.

**Figure 1.**
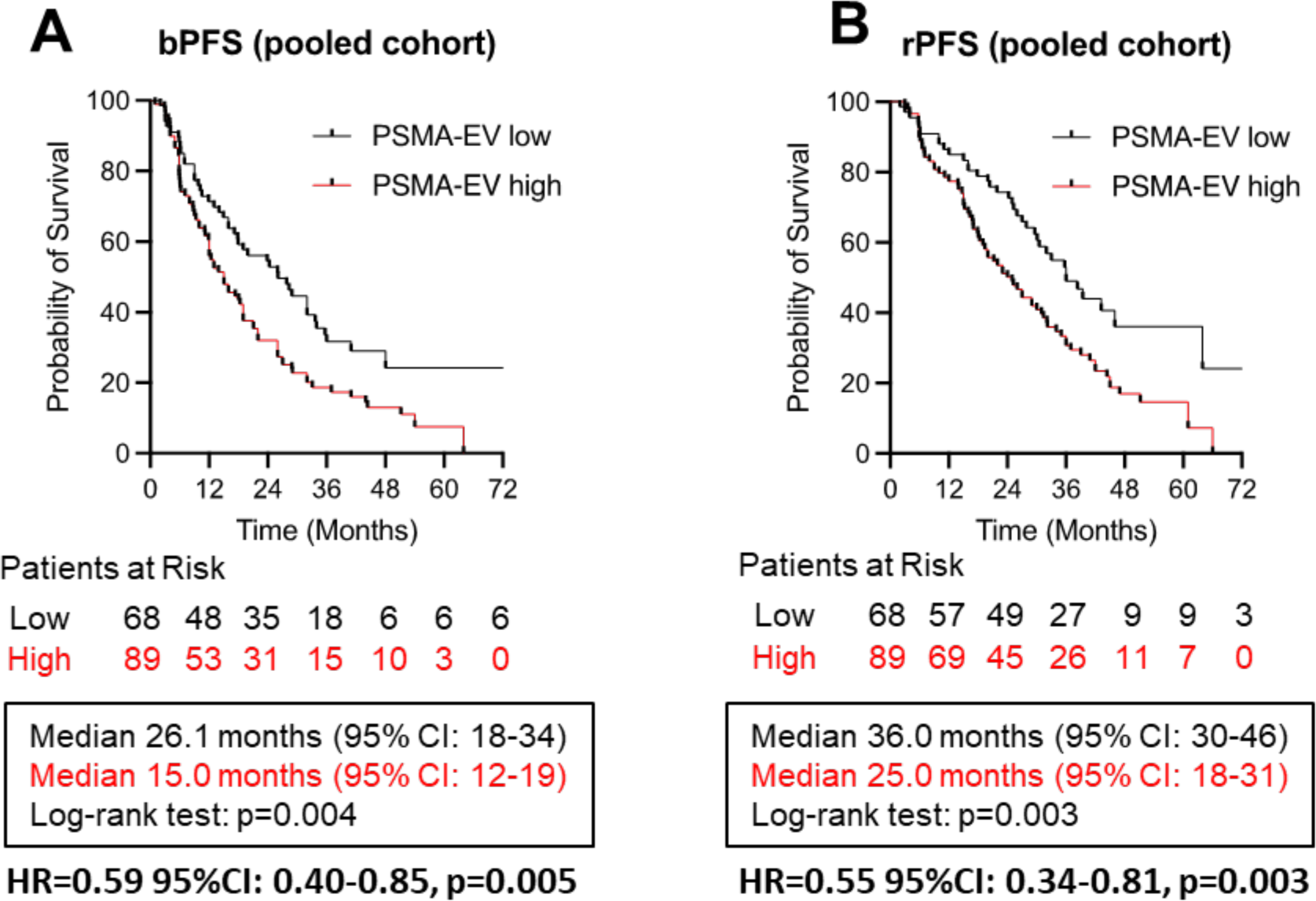
Progression-free survival stratified by baseline PSMA^+^EV levels. Kaplan-Meier curves for biochemical progression-free survival (bPFS) and radiographic progression-free survival (rPFS) in the pooled cohorts.

PSMA^+^EVs remained an independent predictor of risk of both bPFS and rPFS when controlling for the effect of PSA levels, number of lesions, and lesion location in multivariate Cox proportional hazard models. In the pooled cohort, low PSMA^+^EVs was also an independent predictor of bPFS (HR=0.59, 95% CI: 0.40-0.85, p=0.005) and radiographic progression (HR=0.55, 95% CI: 0.34-0.81, p=0.003). (**Supplementary Table S1**).

### Combination baseline PSA and PSMA^+^EVs and oncological outcomes

While patient stratification on PSMA^+^EVs alone produced significant differences in PFS, combining PSMA^+^ EVs and PSA resulted in identification of long-term responders to SABR (**Figure 1A-1B**). Patients were stratified as PSA low/PSMA^+^EV low, PSA high/PSMA^+^EV high, PSA low/PSMA^+^EV high and PSA high/PSMA^+^EV low. In the pooled cohort, 15% (19/127) of patients presented with PSA low/PSMA^+^EV low levels. Median bPFS and rPFS were not reached and PSA low/PSMA^+^EV low was a superior prognostic marker of bPFS and rPFS (**Figure 2A-2B**). Combination of baseline levels of PSMA^+^EV and PSA was associated with lower risk of bPFS (HR=0.34, 95% CI: 0.18-0.58, p=0.0002) and rPFS (HR=0.22, 95% CI: 0.09-0.44, p=0.0001). Progression-free survival stratified by PSMA^+^EV and combination PSMA^+^EV and PSA for the individual cohorts can be seen in **Supplementary Figures S8 and S9**

**Figure 2.**
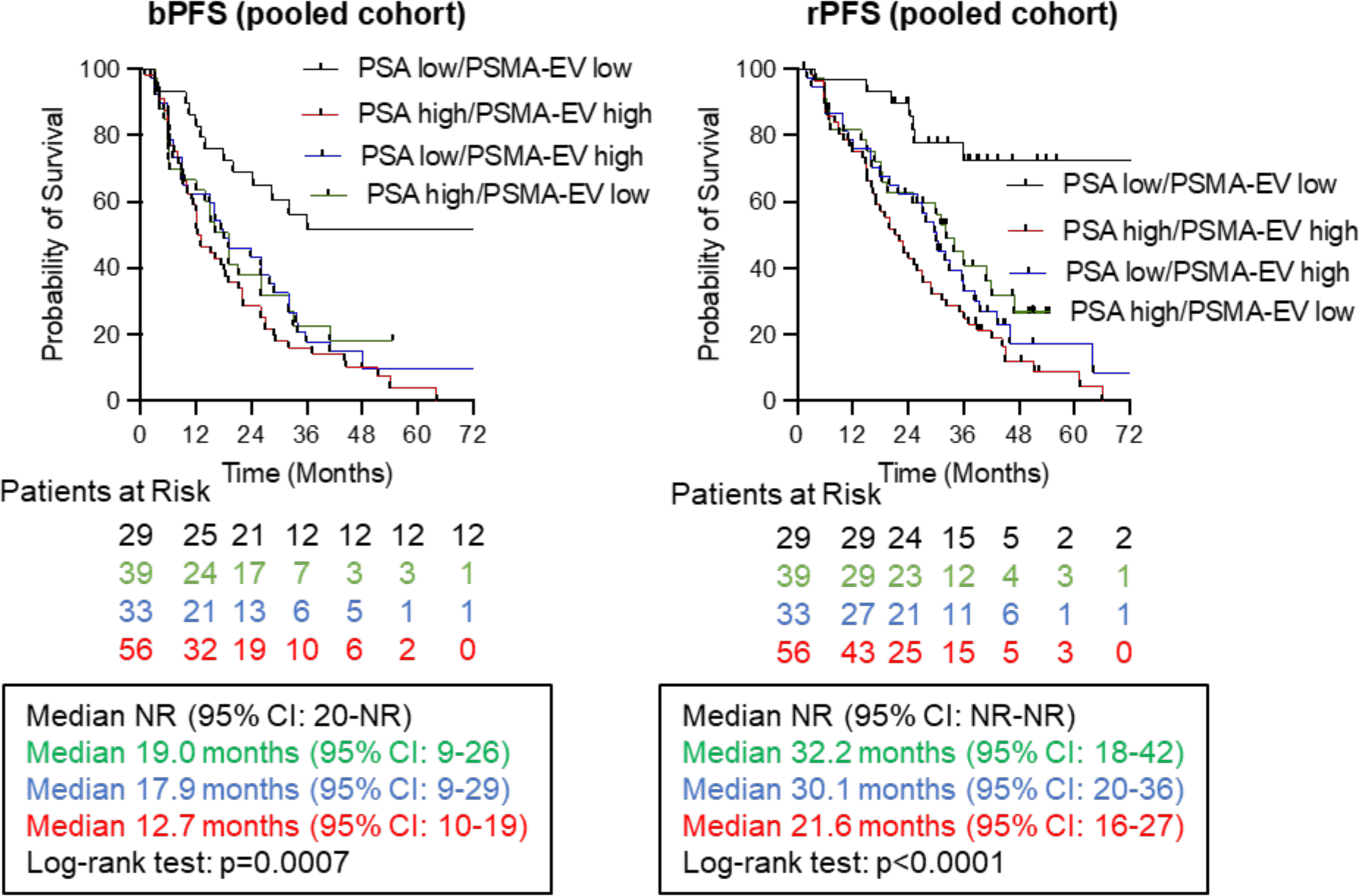
Progression-free survival stratified by baseline PSA and PSMA+EV levels. Kaplan-Meier curves for biochemical progression-free survival (bPFS) and radiographic progression-free survival (rPFS) in the (A-B) the pooled cohorts

### PSMA+EV is a predictive biomarker of response to SABR

We analyzed the baseline levels of PSMA^+^EVs in both SABR and observation arms of the ORIOLE cohort to evaluate their predictive value. Patients who presented with low levels of PSMA^+^EVs show benefit from SABR compared to patients in the observation arm (**Figure 3A)**. Median bPFS was 24.3 and 5.8 months for SABR and observation arms respectively (p=0.003). Patients treated with SABR had a significant lower risk of biochemical progression (HR = 0.19, 95% CI: 0.065-0.644, p=0.004). In contrast, SABR did not show any benefit for patients with high baseline levels of PSMA^+^EVs compared to observation (5.9 and 7.1 months, p=0.95) (**Figure 3B)**. Risk of biochemical progression was not statistically different between both groups (HR = 1.17, 95% CI: 0.498-2.766, p=0.715). Baseline levels of PSMA^+^EVs did not significantly affect risk of radiographic progression (**Supplementary Figure S10**). There was not a significant treatment effect neither within the high PSMA group (HR = 1.39, 95% CI: 0.50-3.84, p=0.512) nor within the low PSMA group (HR = 0.57, 95% CI: 0.14-2.22, p= 0.419).

**Figure 3.**
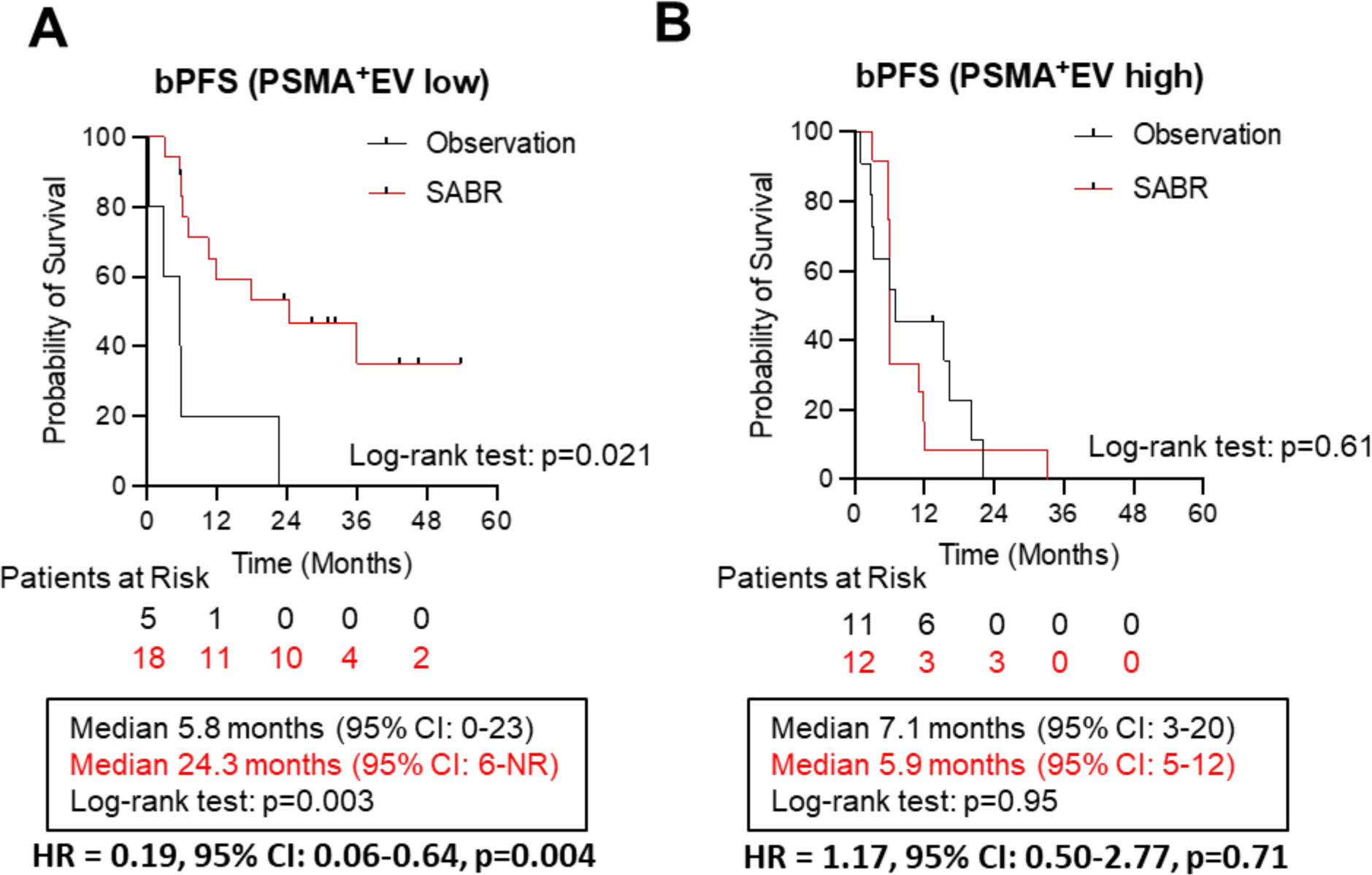
Biochemical progression-free survival stratified by baseline PSMA^+^EV levels in the observation and SABR arms of the ORIOLE trial. Kaplan-Meier curves for biochemical progression-free survival (bPFS) in patients of the ORIOLE SABR and observations arms stratified by baseline levels of PSMA^+^EVs low (A) and high (B).

## Discussion

Systemic therapy with ADT alone or in combination with next generation AR targeted therapy is the standard of care for patients with oligorecurrent CSPC. The benefit of systemic therapy intensification with chemotherapy remains very modest in patients presenting with metachronous low-volume disease (≤ 3 non-visceral metastases).^11^ The concept of metastasis-directed therapy (MDT) with SABR or surgery in oligorecurrent prostate cancer is gaining traction following the positive results from the STOMP and ORIOLE randomized trials.^3,4^ MDT represents a safe and effective treatment option to eliminate visible metastases on imaging without using systemic therapy. Of note, ∼10-15% of patients achieve prolonged disease control with no sign of recurrence on imaging for at least 4 years post-treatment.^12-16^ While local control remains excellent (∼90% at 2 years follow-up) after SABR treatment, oligoprogression remains the major cause of clinical failure post-SABR and it suggests that patients may present at diagnosis with micrometastatic disease below the threshold of detection of both conventional and advanced imaging^17^. To date, treatment with MDT and/or systemic intensified hormonal therapy for patients with oligorecurrent CSPC is informed by clinical presentation such as volume and number of metastases, metastasis location, patient comorbidities and physician experience. The integration of tumor biomarkers with imaging variables better supports treatment decisions and estimation response to SABR.^12^ Our current work demonstrates the clinical value of a pre-treatment PSMA^+^EV liquid biopsy as a non-invasive biomarker to redefine a true oligometastatic disease state and better select patients for SABR.

In this study we assessed 3 potential prognostic biomarkers to SABR; PSMA^+^EVs, PSA, and PSA doubling time in a large multi-institutional dataset of 157 patients with omCSPC. We found that high baseline concentrations of PSMA^+^EVs was a prognostic biomarker of both biochemical and radiographic progression following SABR treatment which is in line with our original proof-of-concept study in omCRPC^6^. The combination of baseline PSA and PSMA^+^EVs was superior to all three individual biomarkers. On assessment with multivariate Cox proportional hazard models, controlling for the effect of PSA levels, number of lesions, and lesion location, PSMA^+^EVs remains an independent prognostic biomarker. In addition, a total of 29/157 (18.5%) patients were identified with PSA low and PSMA^+^EV low and this combination was associated with long PFS. Similar to STOMP and ORIOLE studies, 30/157 patients (19.1%) did not experience either biochemical or radiographic progression at a median follow-up of 40.3 months (95% CI: 36.2-48.5). From those, 16 patients (53.3%) were classified as pre-SABR PSA low and PSMA^+^EV low. Conversely, 109/157 patients (69.4%) showed evidence of radiographic progression at a median follow-up of 20.4 months (95% CI: 17.0-25.30) and only 7 of them (6.4%) had both PSA low and PSMA^+^EV low. Our work demonstrates that pre-SABR blood levels of PSA and PSMA^+^EVs is a blood-based biomarker that can identify patients with truly omCSPC from those who likely harbor micrometastatic disease at diagnosis and may be predictive of response to SABR alone.

We identified PSMA^+^EVs as a predictive biomarker of bPFS in response to SABR from the ORIOLE trial. PSMA^+^EVs was not found to be a predictive biomarker of rPFS likely because of the limited sample size and crossover which occurred in almost all observations arm patients resulting in low differential PFS events. SABR demonstrated durable benefit in patients with PSMA^+^EV low, yet SABR did not provide any benefit in patients with high PSMA^+^EVs compared to observation. Validation of PSMA^+^EVs as a predictive biomarker of both biochemical and radiographic progression is warranted and would provide the first blood-based predictive biomarker for MDT in omCSPC. Moving forward, the future management of omCSPC may be improved where treatment decisions are better informed by advanced PET imaging, blood-based measurement of tumor burden and molecular profiling. We propose that this management approach for oligometastatic prostate cancer should be evaluated in a prospective clinical trial that stratifies treatment based on PSMA^+^EVs and PSA levels similar to the escalation/de-escalation design of the PREDICT-RT trial (NCT04513717).

This study is not without limitations. While the study had access to patient plasma samples from the ORIOLE trial, most of patients in the STOMP trial had stored serum samples. Plasma samples are more suitable than serum for nanoscale flow cytometry-based EV measurement (data not shown). Therefore, we utilized plasma samples from patients recruited outside the STOMP trial but under the same protocol. In line with this, we acknowledge the potential for bias and variability in collection from a non-randomized cohort. Additionally, our study did not consider the somatic mutational profile of patients. Deek et al, found several somatic mutations (e.g TP53, ATM, RB1, BRCA1/2) in omCSPC^18^ that are associated with higher risk of clinical failure following SABR.^12^ In the future, the definition of oligometastasis may not solely rely on the number of metastases but also on the intrinsic tumor biology for which our PSMA^+^EV assay does not currently assess. Further studies leveraging EV’s molecular cargo may provide further utility to EVs as liquid biopsy.

We found median levels of PSMA^+^EVs significantly lower in the ORIOLE cohort compared to the STOMP-like cohort which is counterintuitive considering that ORIOLE trial enrolled patients diagnosed with conventional imaging. We cannot exclude variability in blood collection, handling and storage conditions which could have impacted PSMA^+^EV measurement^19^ including freeze-thaw cycles (unpublished data).

## Conclusion

In conclusion, PSMA^+^EVs is a novel prognostic biomarker of tumor burden and micrometastatic disease in omCSPC and a predictive biomarker of biochemical progression for SABR in omCSPC. Oligometastatic prostate cancer represents a heterogenous patient population and the combination of PSMA^+^EV and PSA can help refine selection of patients who will experience durable disease-free survival in response to SABR. This observational study provides the first clinical use of extracellular vesicles on a prospective trial of omCSPC and strengthens the clinical value of PSMA^+^EVs for personalized radiation therapy. The notable divergence in survival curves stratified by PSMA+EV and PSA levels highlights the need for prospective validation of PSMA^+^EVs as a predictive biomarker for SABR. Going forward, biomarker-directed risk stratification and personalization tested in prospective clinical trial protocols is needed.

## Data availability statement

Individual deidentified patient data related to the results reported in this article will be shared as will the individual study protocols. The data will become available beginning 1 year and for 3 years following publication to researchers with a methodologically sound proposal to achieve aims in the previously said sound proposal. Proposals should be directed toward the corresponding authors Dr. Fabrice Lucien and Sean Park or the principal investigators of the study cohorts Drs Piet Ost, Carole Mercier and Phuoc T. Tran.

## Funding statement

This work was supported by the Erivan K. and Helga Haub Family Fund in Image-Guided Urology (FL).

## Disclosure statement

Innovations and intellectual property generated during this research have been protected under a patent application filed with the help of Mayo Clinic Ventures. JJO - Research support from Prostate Cancer Foundation, Nanotics, and Genentech. Honoraria or Travel Support from Nanotics and Partner Therapeutics. FL – Research support, Consulting and Travel Support from Nanotics, Scientific Advisory board for Mursla Bio, Licensing Agreement and Royalties from Early is Good. PTT – Research support from Astellas, Bayer Healthcare, and RefleXion Medical Inc; Personal fees from Bayer Healthcare, RefleXion, Pfizer, Lantheus, Janssen-Taris Biomedical, Regeneron, Pfizer and AstraZeneca; and has a patent 9114158 - Compounds and Methods of Use in Ablative Radiotherapy licensed to Natsar Pharm.. DSC – Research support from Janssen. Honoraria, consulting, and/or travel from Novartis (to institution), Janssen (to institution), Targeted Oncology, IntrinsiQ, MJH Life Sciences, International Centers for Precision Oncology Foundation, Prostate Cancer Foundation.

## Ethics approval statement

This work was approved by the Mayo Clinic Institutional Review Board with informed patient consent under IRB 21-004451.

## Translational Relevance

While metastasis directed therapy is a novel and a potetnially curative treatment option for oligometatastic prostate cancer, the 2 prospective clincal trials in this setting have demostrated mixed outcomes. While some patients demonstrate a durable response, many patietns have a limited response. Using samples from the ORIOLE Trial and the STOMP Trial protocol, we have evelautated PSMA+ extracellular vesicles as a potenital biomaker of resposne. Out data presented here represents the first blood based biomarker in prostate cancer that is both a prognostic biomarker and predicitve of response to metastasis directed therapy. This biomarker allows for appropriate patient selection for those most likely to benefit from metastasis directed therapy and to by passs metastasis directed therapy in those unlikely to benefit. This has immense clinical implications and will help expand the role of metastisis directed therapy.

## Supplements

**Supplementary Figure S1. Diagram of the study cohorts**

**Supplemental Figure S2. Oncological outcomes of the study cohorts**

Kaplan-Meier curves for (A) biochemical progression-free survival (bPFS) and (B) radiographic progression-free survival (rPFS) in the Ghent U, Iridium Net, ORIOLE cohorts.

**Supplemental Figure S3. Comparative analysis of baseline PSA and PSMA+EV between study cohorts**

Baseline levels of (A) PSA and (B) PSMA+EV in the three cohorts. P values were determined by Kruskal-Wallis test. Spearman coefficients and P value for correlation between baseline levels of PSA and PSMA+EV for the three cohorts are presented in C.

**Supplemental Figure S4. Progression-free survival stratified by number of lesions treated**

Kaplan-Meier curves for biochemical progression-free survival (bPFS) and radiographic progression-free survival (rPFS) in (A-B) the ORIOLE cohort and (C-D) the GhentU/Iridium Network pooled cohort.

**Supplemental Figure S5. Progression-free survival stratified by baseline PSA levels in pooled cohorts**

Kaplan-Meier curves for biochemical progression-free survival (bPFS) and radiographic progression-free survival (rPFS) in the pooled cohort

**Supplemental Figure S6. Progression-free survival stratified by baseline PSA levels in separate study cohorts**

Kaplan-Meier curves for biochemical progression-free survival (bPFS) and radiographic progression-free survival (rPFS) in (A-B) the ORIOLE cohort and (C-D) the GhentU/Iridium Network pooled cohort

**Supplemental Figure S7. Progression-free survival stratified by baseline PSA DT levels**

Kaplan-Meier curves for biochemical progression-free survival (bPFS) and radiographic progression-free survival (rPFS) in (A-B) the ORIOLE cohort and (C-D) the GhentU/Iridium Network pooled cohort.

**Supplemental Figure S8. Progression-free survival stratified by PSMA+EV levels**

Kaplan-Meier curves for biochemical progression-free survival (bPFS) and radiographic progression-free survival (rPFS) in (A-B) the ORIOLE cohort and (C-D) the GhentU/Iridium Network pooled cohort.

**Supplemental Figure S9. Progression-free survival stratified by baseline PSA and PSMA^+^EVs**

Kaplan-Meier curves for biochemical progression-free survival (bPFS) and radiographic progression-free survival (rPFS) in (A-B) the ORIOLE cohort and (C-D) the GhentU/Iridium Network pooled cohort.

**Supplementary Figure S10. Association of baseline PSMA^+^EV levels with radiographical PFS in ORIOLE SABR and observation arms**

Kaplan-Meier curves for radiopgrahic progression-free survival (rPFS) in patients of the ORIOLE SABR and observations arms stratified by baseline levels of PSMA^+^EVs low (A) and high (B).

## Supplementary Tables

**Table S1.** Multivariate analysis of the risk of progression for baseline PSMA^+^EV, PSA and PSA DT in the pooled STOMP-like and ORIOLE cohort.

|  |  |
| --- | --- |
| Number of Observations Read | 157 |
| Number of Observations Used | 145 |

bPFS
| Parameter |  | Parameter Estimate | Standard Error | Chi-Square | Pr > ChiSq | Hazard Ratio | Reference |
| --- | --- | --- | --- | --- | --- | --- | --- |
| Number of lesions | 2 | -0.25805 | 0.25075 | 1.0591 | 0.3034 | 0.773 | Number of lesions: 1 |
| Number of lesions | 3 | 0.07459 | 0.26628 | 0.0785 | 0.7794 | 1.077 | Number of lesions: 1 |
| Number of lesions | 4+ | -0.17028 | 0.39384 | 0.1869 | 0.6655 | 0.843 | Number of lesions: 1 |
| Lesion location | bone only | 0.22399 | 0.33368 | 0.4506 | 0.5021 | 1.251 | Lesion location: mixed |
| Lesion location | node only | -0.48574 | 0.31516 | 2.3755 | 0.1232 | 0.615 | Lesion location: mixed |
| Baseline PSA | Low | -0.38172 | 0.20972 | 3.3128 | 0.0687 | 0.683 | High PSA |
| PSA DT | ≤ 3 months | 0.25414 | 0.21213 | 1.4353 | 0.2309 | 1.289 | PSA DT > 3 months |
| Baseline PSMA<sup>+</sup>EV | Low | **-0.53541** | **0.21288** | **6.3254** | **0.0119** | **0.585** | **High PSMA<sup>+</sup>EV** |

rPFS
| Parameter |  | Parameter Estimate | Standard Error | Chi-Square | Pr > ChiSq | Hazard Ratio | Label |
| --- | --- | --- | --- | --- | --- | --- | --- |
| Number of lesions | 2 | -0.0008238 | 0.26213 | 0 | 0.9975 | 0.999 | Number of lesions: 1 |
| Number of lesions | 3 | 0.13592 | 0.28534 | 0.2269 | 0.6338 | 1.146 | Number of lesions: 1 |
| Number of lesions | 4+ | 0.30743 | 0.42431 | 0.525 | 0.4687 | 1.36 | Number of lesions: 1 |
| Lesion location | bone only | 0.08553 | 0.34717 | 0.0607 | 0.8054 | 1.089 | Lesion location: mixed |
| Lesion location | node only | -0.40535 | 0.3215 | 1.5896 | 0.2074 | 0.667 | Lesion location: mixed |
| Baseline PSA | Low | -0.58205 | 0.23418 | 6.1774 | 0.0129 | 0.559 | High PSA |
| PSA DT | ≤ 3 months | 0.21408 | 0.23034 | 0.8638 | 0.3527 | 1.239 | PSA DT > 3 months |
| Baseline PSMA<sup>+</sup>EV | Low | **-0.60386** | **0.23196** | **6.7774** | **0.0092** | **0.547** | **High PSMA<sup>+</sup>EV** |

