## Supplementary Materials for "PSMA+ Extracellular Vesicles are a Biomarker for SABR in Oligorecurrent Prostate Cancer Analysis from the STOMP-like and ORIOLE trial cohorts"

### Figure S1. Diagram of the study cohorts

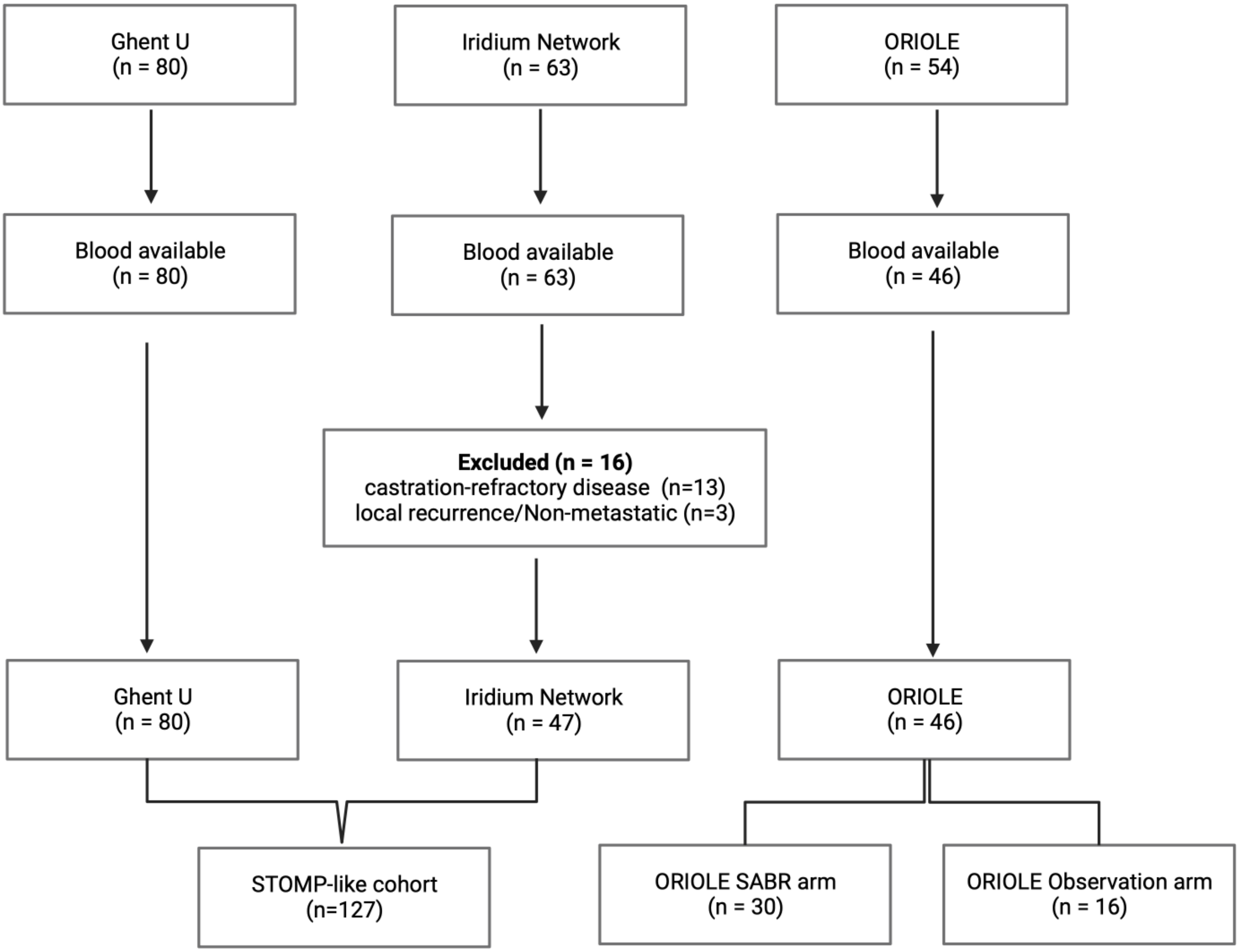

### Figure S2. Oncological outcomes for the study cohorts

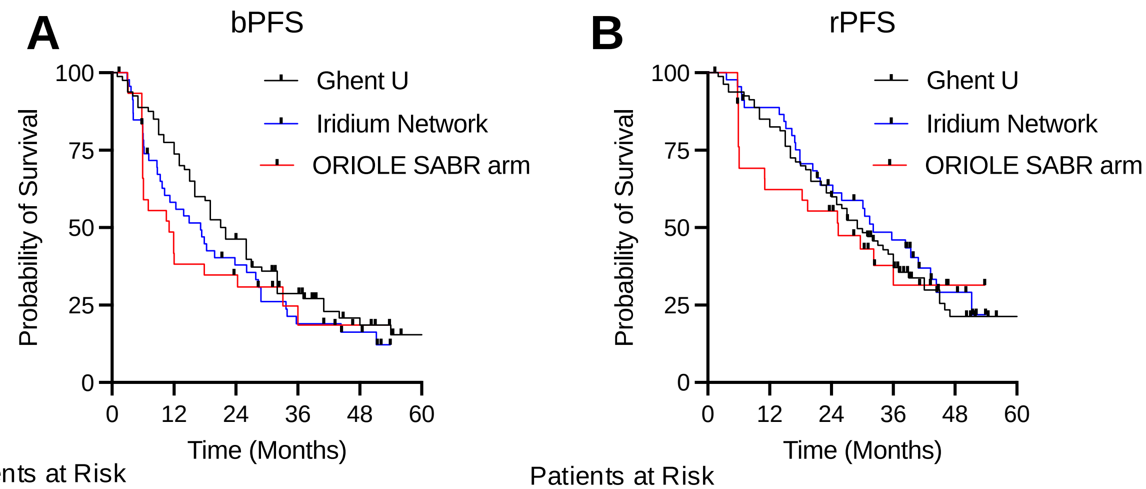


**A**

Patients at Risk

| | | | | | | |
|---|---|---|---|---|---|---|
| Ghent U | 80 | 62 | 37 | 25 | 9 | 4 |
| Iridium Net | 47 | 27 | 18 | 9 | 6 | 1 |
| ORIOLE | 30 | 12 | 10 | 4 | 2 | 1 |

Median 21.5 months
Median 17.2 months
Median 11.1 months

**B**

Patients at Risk

| | | | | | | |
|---|---|---|---|---|---|---|
| Ghent U | 80 | 68 | 49 | 29 | 11 | 6 |
| Iridium Net | 47 | 40 | 27 | 19 | 7 | 1 |
| ORIOLE | 30 | 19 | 16 | 6 | 2 | 1 |

Median 29.0 months
Median 32.1 months
Median 25.3 months

### Figure S3. Comparative analysis of baseline PSA and PSMA+EV between study cohorts

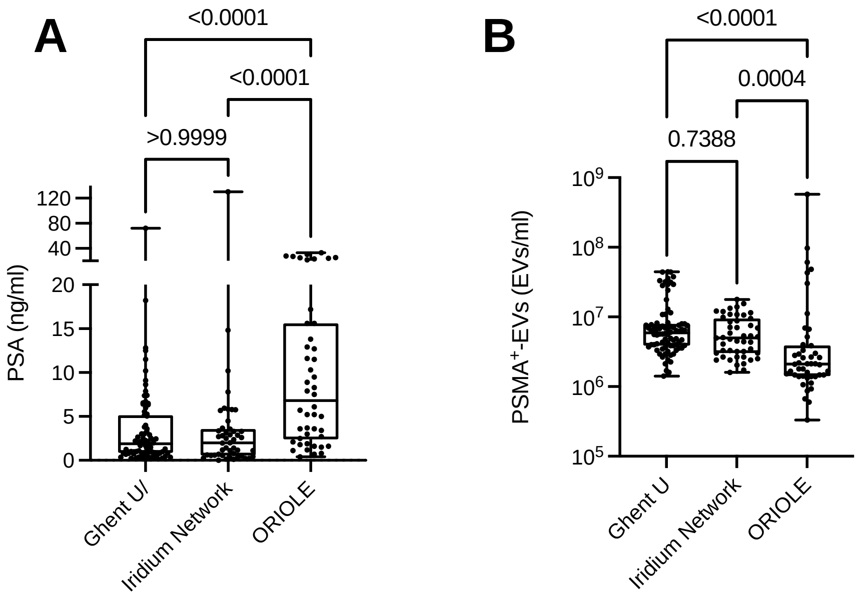


C

| | Spearman r (95% CI) | P value |
|---|---|---|
| Ghent U | -0.10 (-0.32-0.12) | 0.38 |
| Iridium Network | 0.001 (-0.29-0.30) | 0.99 |
| ORIOLE | -0.07 (-0.43-0.31) | 0.72 |

### Figure S4. Progression-free survival stratified by number of lesions treated

**A**

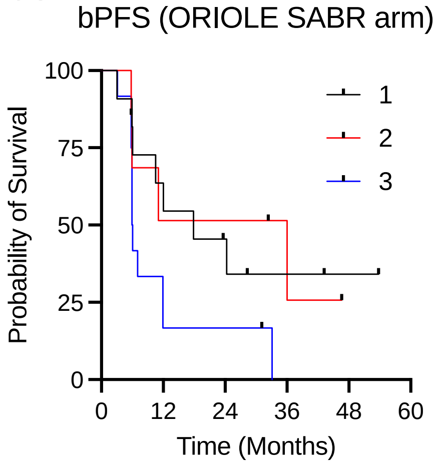


Patients at Risk

| | | | | |
|---|---|---|---|---|
| 11 | 7 | 5 | 3 | 2 |
| 7 | 4 | 4 | 2 | 1 |
| 12 | 4 | 4 | 0 | 0 |

Median 17.9 months
Median 36.0 months
Median 6.0 months
Log-rank test: p=0.10

**B**

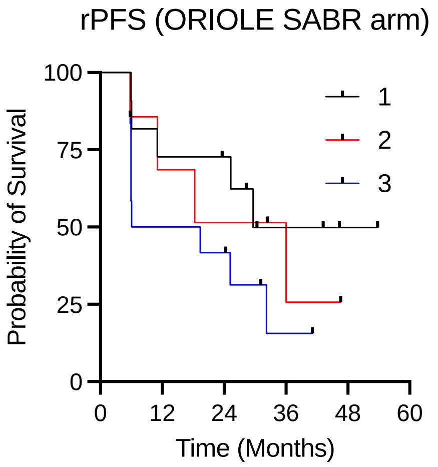


Patients at Risk

| | | | | |
|---|---|---|---|---|
| 11 | 9 | 8 | 4 | 2 |
| 7 | 5 | 4 | 2 | 1 |
| 12 | 7 | 6 | 2 | 1 |

Median 29.6 months
Median 36.0 months
Median 12.8 months
Log-rank test: p=0.24

**C**

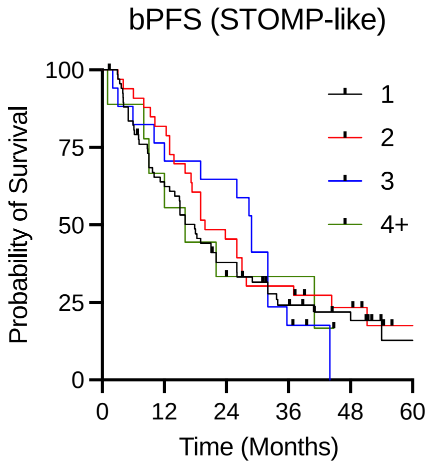


Patients at Risk

| | | | | | |
|---|---|---|---|---|---|
| 68 | 42 | 26 | 14 | 8 | 3 |
| 33 | 28 | 16 | 11 | 7 | 2 |
| 17 | 13 | 12 | 7 | 0 | 0 |
| 9 | 6 | 3 | 3 | 1 | 1 |

Median 17.9 months
Median 19.9 months
Median 28.9 months
Median 16 months
Log-rank test: p=0.90

**D**

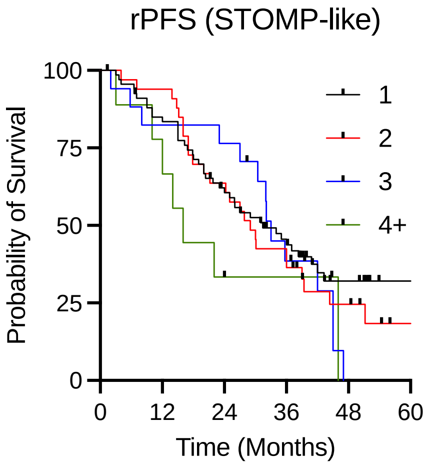


Patients at Risk

| | | | | | |
|---|---|---|---|---|---|
| 68 | 56 | 39 | 25 | 11 | 5 |
| 33 | 32 | 22 | 14 | 7 | 2 |
| 17 | 15 | 14 | 7 | 0 | 0 |
| 9 | 7 | 3 | 3 | 0 | 0 |

Median 31.4 months
Median 29.0 months
Median 33.0 months
Median 16.0 months
Log-rank test: p=0.47

### Figure S5. Progression-free survival stratified by baseline PSA levels in pooled cohorts

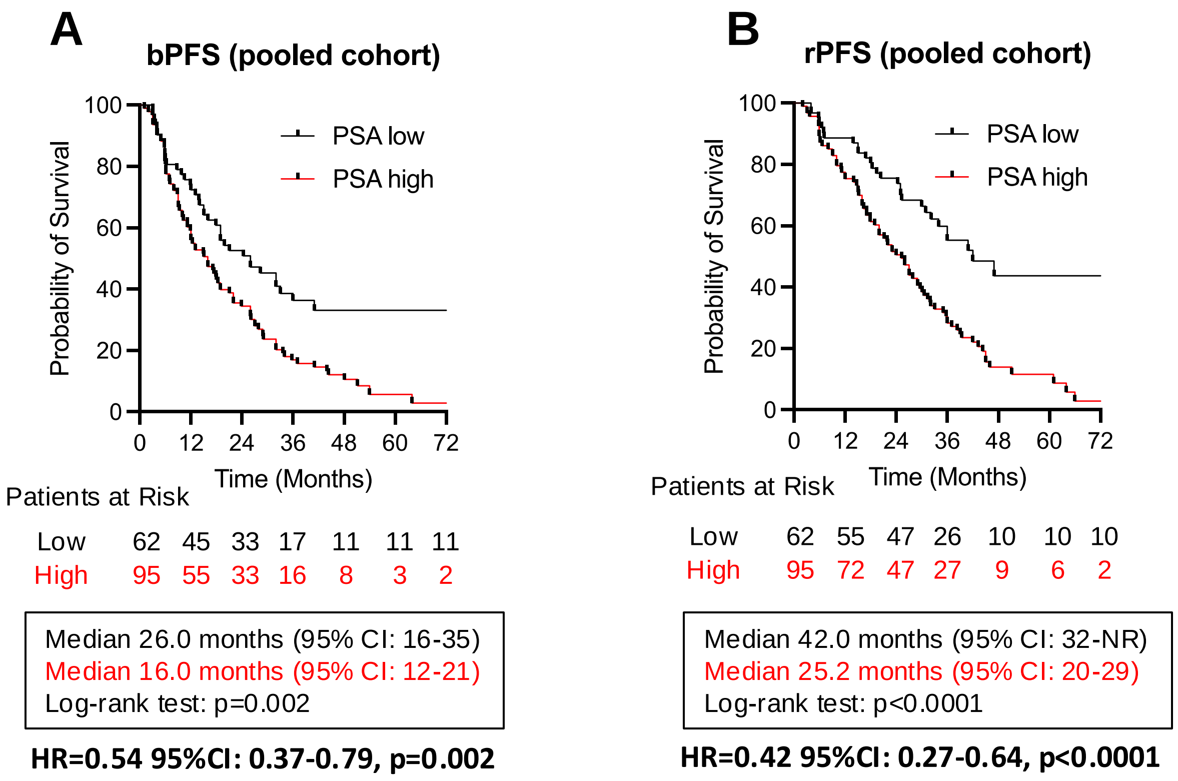

### Figure S6. Progression-free survival stratified by baseline PSA levels in separate study cohorts

**A**

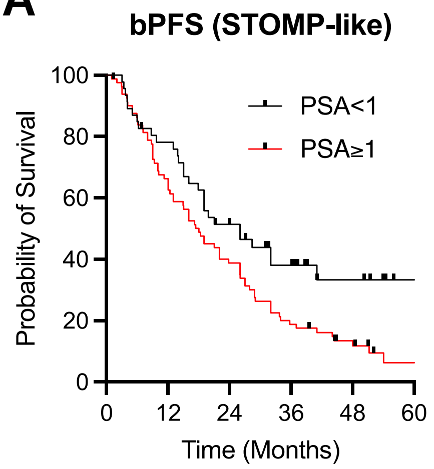


Patients at Risk

| | | | | | | |
|---|---|---|---|---|---|---|
| Low | 46 | 36 | 22 | 15 | 7 | 2 |
| High | 81 | 53 | 32 | 16 | 8 | 3 |

Median 26.0 months
Median 18.0 months
Log-rank test: p=0.011

**B**

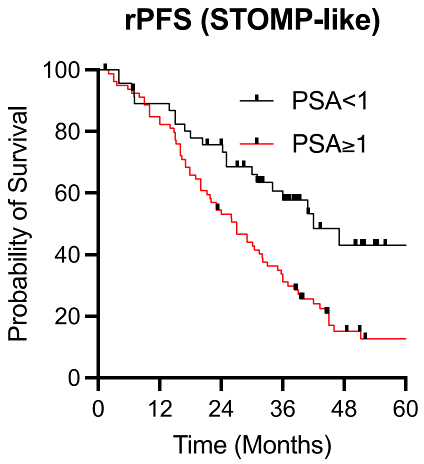


Patients at Risk

| | | | | | | |
|---|---|---|---|---|---|---|
| Low | 46 | 41 | 33 | 21 | 9 | 2 |
| High | 81 | 67 | 42 | 26 | 9 | 5 |

Median 42.0 months
Median 27.0 months
Log-rank test: p=0.009

**C**

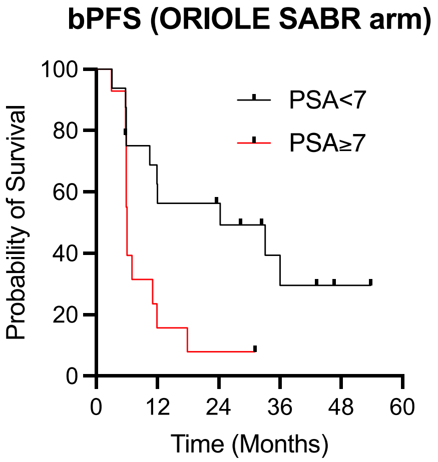


Patients at Risk

| | | | | | |
|---|---|---|---|---|---|
| Low | 16 | 10 | 9 | 4 | 2 |
| High | 14 | 3 | 2 | 0 | 0 |

Median 24.3 months
Median 6.1 months
Log-rank test: p=0.009

**D**

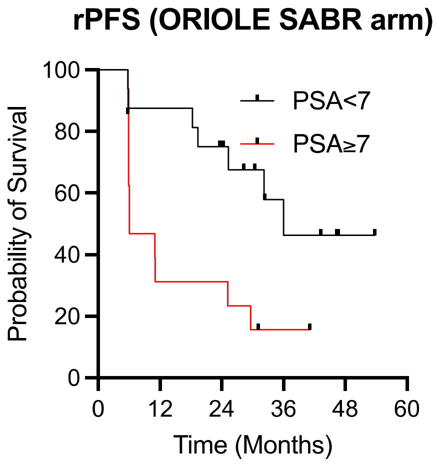


Patients at Risk

| | | | | | |
|---|---|---|---|---|---|
| Low | 16 | 15 | 12 | 5 | 2 |
| High | 14 | 5 | 5 | 2 | 0 |

Median 36.0 months
Median 6.1 months
Log-rank test: p=0.0073

### Figure S7. Progression-free survival stratified by baseline PSA doubling time (PSA DT)

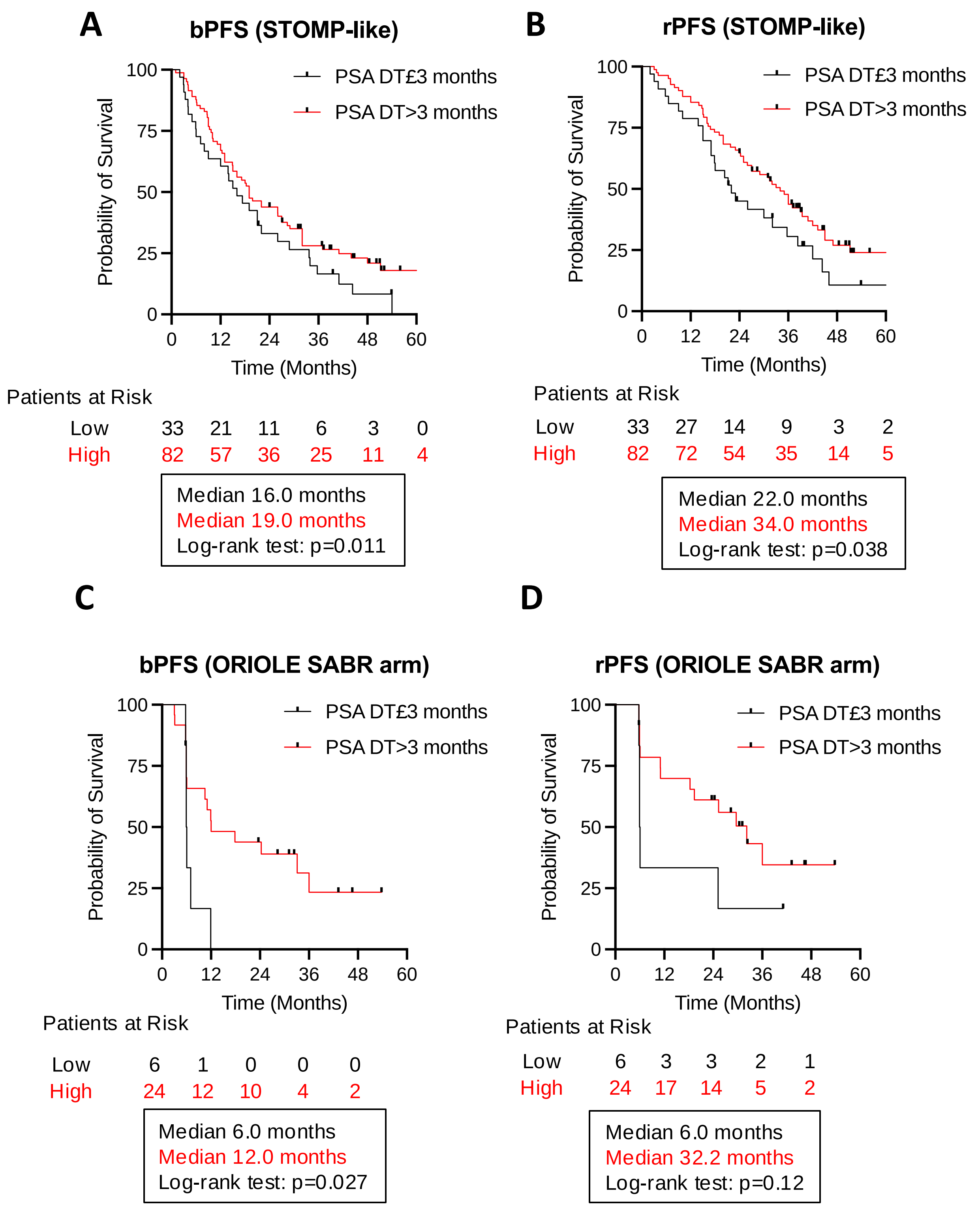

### Figure S8. Progression-free survival stratified by baseline PSMA⁺EV levels

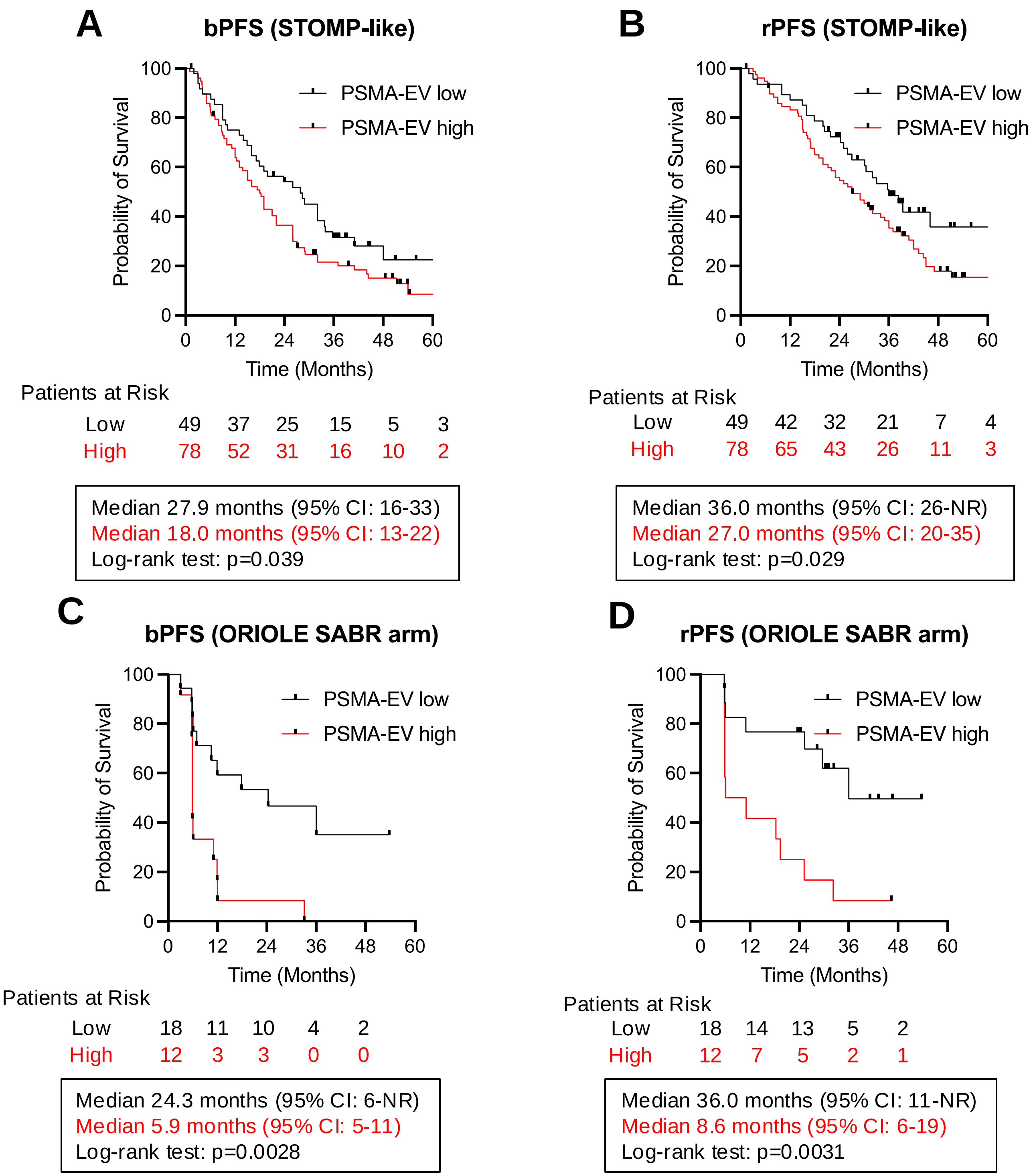

### Figure S9. Progression-free survival stratified by baseline PSA and PSMA⁺EVs

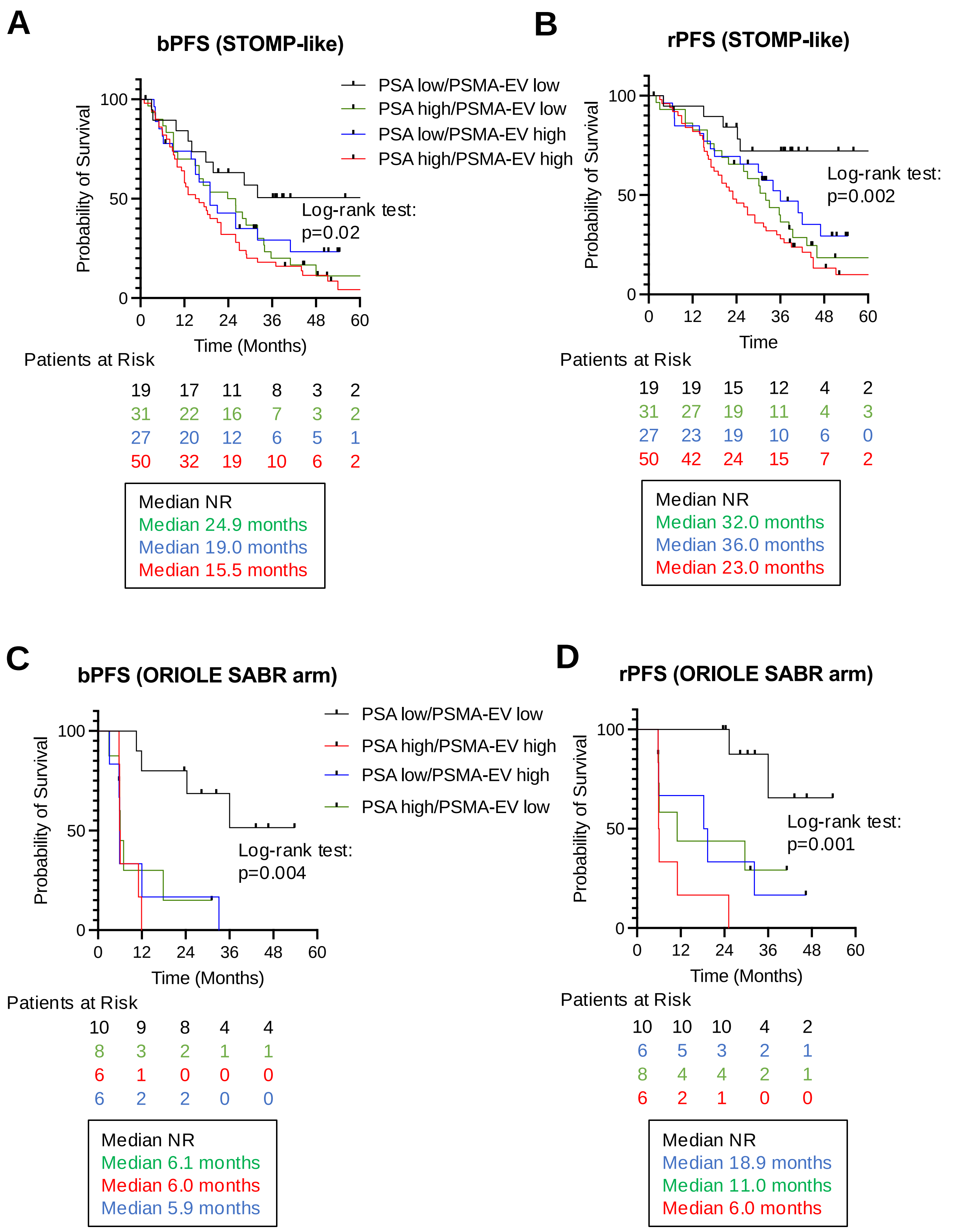

### Figure S10. Association of baseline PSMA$^{+}$EV levels with radiographical PFS in ORIOLE SABR and observation arms

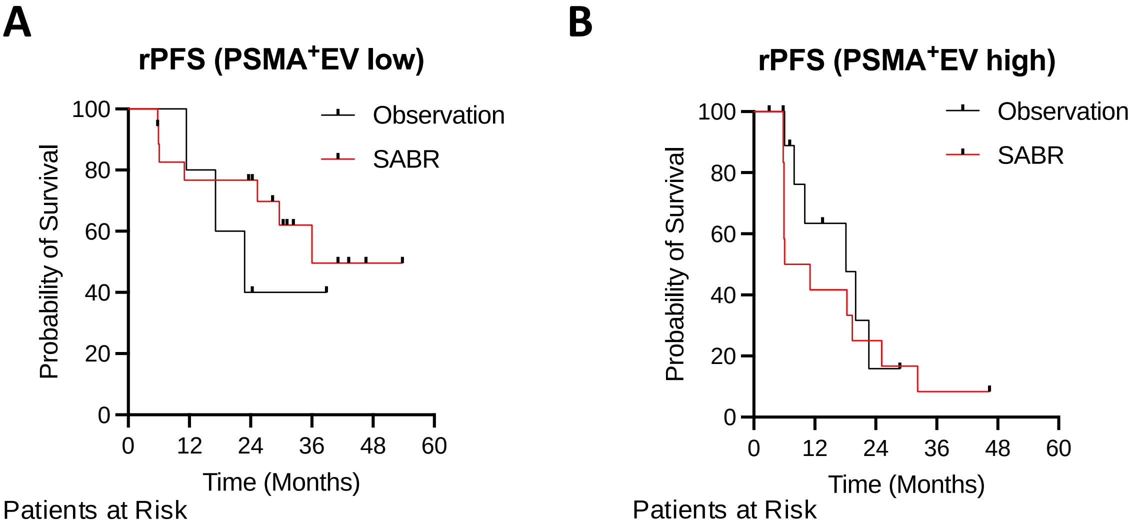


**A** — Patients at Risk

| | | | | |
|---|---|---|---|---|
| 5 | 5 | 2 | 1 | 1 |
| 18 | 14 | 13 | 5 | 2 |

Median 22.8 months
Median 36.0 months
Log-rank test: p=0.43

**B** — Patients at Risk

| | | | | |
|---|---|---|---|---|
| 11 | 6 | 2 | 1 | 0 |
| 12 | 3 | 3 | 0 | 0 |

Median 18.1 months
Median 8.6 months
Log-rank test: p=0.49
